# Understanding COVID-19 Vaccine Hesitancy Among Black and Afro-Latinx Pregnant Individuals: A Mixed-Methods Approach Utilizing Focus Groups and Social Media Ad Reaction

**DOI:** 10.64898/2026.01.06.25343048

**Authors:** Elizabeth Cox, Emma Every, Rachel Johnson, Melissa Baker, Magali Sanchez, Isabelle Crary, Carly Baxter, Simone Stapley, Jeff Munson, Alex Stonehill, Kristina M. Adams Waldorf

## Abstract

Pregnant individuals have a greater susceptibility to severe disease from the coronavirus 2019 disease (COVID-19). Pregnant people also tend to be vaccine-hesitant, which is even more pronounced in certain racial and ethnic minority groups. The study objective was to determine whether social media ads promoting COVID-19 vaccination could influence vaccination likelihood among pregnant and recently pregnant participants who self-identified as Black or African American. Participants were interviewed individually or in focus groups to explore their attitudes about vaccination and to ask them to rate their COVID-19 vaccination likelihood after seeing a panel of ads featuring different messengers (e.g., doctor, peer, elder, faith leader) and content types (e.g., social proof, text-heavy, fear-based, activation). Ad ratings were analyzed using linear mixed models to examine the effect of vaccination status, ad messenger, and ad content type. Interviews were coded and analyzed for qualitative themes. Ad scores differed significantly by vaccine status, with vaccinated participants rating ads as more likely to inspire vaccine uptake, while unvaccinated participants rated ads negatively. No specific messenger or content type was rated as more probable to motivate vaccination. There was a significant interaction between faith-based messengers and COVID-19 vaccination status, with faith leaders perceived as more favorable by unvaccinated participants (p=0.008). Vaccine-hesitant respondents cited mistrust of healthcare providers and fears of medical racism. Although we did not identify content types that might be helpful in a public health vaccine campaign targeting Black pregnant people, faith leaders may be a trusted messenger for unvaccinated individuals.

## Introduction

The coronavirus 2019 disease (COVID-19) pandemic was associated with vaccine disinformation on social media and other platforms suggesting adverse effects of vaccination on women’s fertility and pregnancy safety.[1–4] As pregnant people had higher rates of morbidity and mortality from COVID-19, vaccine disinformation threatened their health by increasing the likelihood that they would forego vaccination.[5–7] Vaccine decision-making in pregnancy is already more complicated than in the non-pregnant state because it involves weighing risks and benefits for the pregnant person and fetus.[8] Unsurprisingly, COVID-19 vaccine uptake in pregnant people lagged vaccination in non-pregnant individuals.[6,9] Public health campaigns leveraging social media can reach millions of people in a cost-effective and rapid manner. Designing effective advertisements promoting vaccination for a vaccine hesitant pregnant population requires knowledge of trusted messengers and content that are embraced by demographic groups.[10] The attitude towards COVID-19 vaccination at the intersection of pregnancy and minority racial/ethnic groups, which are historically under-vaccinated, is poorly understood.

Factors contributing to vaccine hesitancy in Black and African American pregnant people are understudied. Potential explanations for vaccine hesitancy in Black pregnant people have included disinformation and inequitable access to vaccination.[6,9] White patients are more likely to have access to a primary care provider and be insured than Black patients.[11][12] Black pregnant people are also more likely to experience maternal mortality, independent of insurance status[13–15]. Finally, historical acts of medical racism (i.e., Tuskegee syphilis study, forced sterilization of Black women) are additional factors that may contribute to a general mistrust of healthcare systems.[13–15]

We have recently studied the ad messengers and content types preferred by pregnant individuals from a rural White and Spanish-Speaking Hispanic racial/ethnic group in the Western U.S.[16,17] In this study, we have used a similar mixed-methods study design to investigate attitudes toward vaccination and how specific messengers and content type in social media ads are perceived by Black pregnant or recently pregnant people.

## 2. Materials and Methods

### Study Design

This was a mixed-methods study designed to capture both qualitative and quantitative data about vaccine hesitancy and reactions to public health advertisements. We gathered qualitative data via direct interviews and focus groups discussing participant experiences with COVID-19 vaccination and pregnancy. Participant inclusion criteria included being currently or recently pregnant (within last 6 months) and self-identification as Black, Afro-Latinx, or mixed race with Black or African American ethnicity. Quantitative data consisted of individual ratings after viewing advertisements promoting COVID-19 vaccination. The study was approved by the University of Washington Institutional Review Board, and we obtained informed consent from all respondents before conducting the interviews.

### Key Informant Interviews

Key informant interviews shaped the development of the interview facilitator guides for the focus groups. We conducted one key informant interview with a physician who provided guidance on experiences specifically with Black and Afro-Latinx pregnant individuals.

### Patient and Public Involvement

Feedback from the key informant interview helped shape our research question and focus group questions. We also vetted the focus group questions, focus group guide, and advertisements with several employees of Black Health, Inc. (formerly the National Black Leadership Commission on Health). Feedback from these interviews shaped the questions and study materials. We also worked with Black Health to identify the method of participant recruitment and format that study interviews would take place in. Focus groups were held during community baby shower events organized by Black Health or their community partners to meet community members in trusted spaces. Dissemination of these research findings will occur through Black Health to their Board and the communities participating in the research.

### Participants and Procedures

Our study population included 113 participants from urban areas across the United States who were at least 18 years of age and were currently pregnant or had been pregnant within 6 months of the study. These urban areas were the Bronx, Brooklyn, Manhattan, Queens, Staten Island, Atlanta, Detroit, Syracuse, and Upstate New York. Black Health recruited participants directly through hosting community “Baby Showers,” and partnering with community organizations with a direct link to Black or African American pregnant or recently pregnant people including the Afiya Center, Mothering Justice, and the New York Police Department Community Engagement Office.

### Measures and Instruments

Study participants completed a 14-question demographic survey in REDCap that included questions on race, socioeconomic status, political and religious affiliations, and COVID-19 vaccination status. Participants then completed a 30–45-minute direct interview or facilitated focus group focused on participants’ feelings toward COVID-19 vaccination and risk, their trusted sources of information, and health decision-making processes during pregnancy.

Finally, participants were shown four sample advertisements promoting the COVID-19 vaccine during pregnancy. They completed an additional REDCap survey that measured their feelings towards the advertisements using a self-reported five-point Likert scale and an open-ended question.

### Ad Design

To determine respondents’ opinions on social media ads’ messengers and content, we created 19 sample social media ads, which tested four types of messengers: peer, doctor, elder, and faith leader, and five types of content: activation, social proof, text heavy, appeal to protect, and information (negative outcomes; Figure 1, enlarged ads shown in Figure S1-S5). A nuanced approach was used to align faith-based messengers with respondents’ religious affiliations, leveraging data collected earlier in the REDCap survey. This personalized strategy triggered branching logic within the advertisement display, ensuring that religious figures such as priests, pastors, or Imams were presented based on participants’ self-identified faiths. For the content types, the ads that appealed to protection conveyed the message that becoming vaccinated against COVID-19 would help to protect the family or the fetus from disease. In the text-heavy ads, the goal was to provide a short, persuasive list of reasons to get vaccinated. Social proof ads conveyed the message that the vaccine had already been safely administered to thousands of other pregnant women. Ads featuring information on adverse outcomes underscored the increased risks in pregnancy for acquiring COVID-19 for unvaccinated women. An activation ad was meant to encourage or “nudge” the participant to get the vaccine, by providing a reminder for someone who has decided to vaccinate but has not followed through. Several combinations of content and messenger type were unrealistic and therefore not used (e.g., faith + social proof). Ads were designed by aligning imagery and linguistic choice with the target audience and were vetted with key informants from Black Health. A deliberate effort was made to prominently feature women and healthcare professionals, mirroring the demographics of the intended audience.

**Figure 1.**
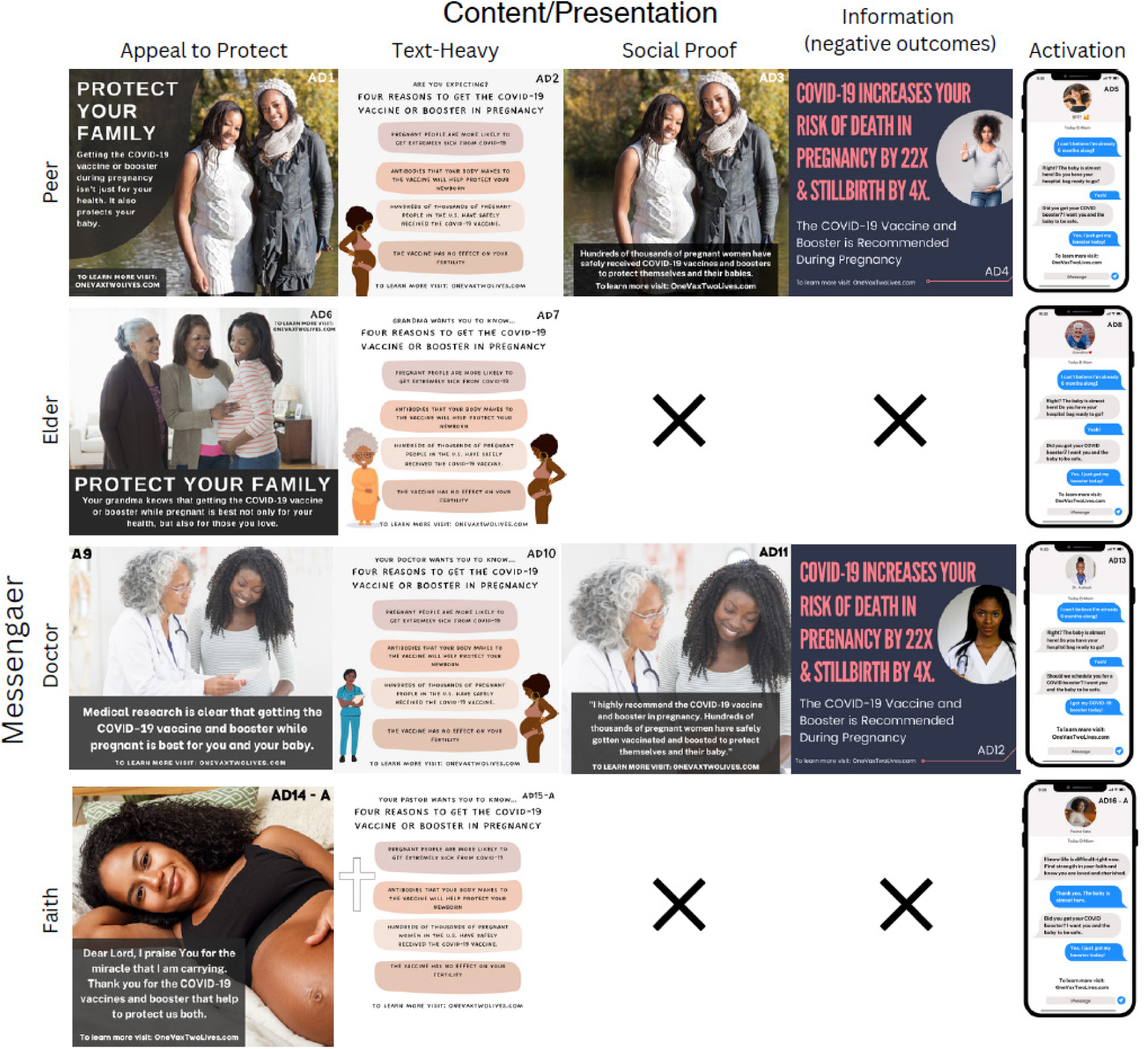
Social Media Ads Shown to Participants. This figure depicts the social media ads shown to participants broken down by messenger and content type. In some cases, combinations of messengers and content were unrealistic (e.g., elder, social proof) and so these ads are indicated by an “X,” as they were not created. Enlarged ads are shown in Supplemental Figures S1-S5.

Ads were randomized into 10 ad sets, each showing 4 ads with different combinations of messenger and content types (Fig. 1, Table S1). To evaluate participants’ reactions to ad messenger and content, we asked participants one qualitative and one quantitative question in response to each ad. First, a qualitative question was asked to determine the initial reaction to each ad, including what the respondent liked and disliked about it. The quantitative question asked respondents to rate each ad on a 1-5 Likert scale of whether they would be more or less likely to receive the COVID-19 vaccine after seeing the ad.

### Data analysis

We used a mixed methods approach to analyze qualitative and quantitative data from direct interviews and focus groups (Table S2). For the qualitative analysis, each interview de-identified and blind coded twice in the software Dedoose using a thematic codebook (Table S3) by two different coders. Following an iterative approach to developing the codebook, codes were initially developed based on existing knowledge, research, and the interview guide, and then revisited and refined to fully and accurately capture emerging themes in the transcripts. Researchers kept notes regarding coding framework and respondents’ perspectives. The codebook was defined before we performed an extensive analysis. We did not compute inter-rater reliability, but coders met after blind-coding to discuss discrepancies. Disagreements over code assignments were rare and resolved through discussion.

For the quantitative analysis, we assessed the effects of messenger type and content type on ad ratings using linear mixed models (R packages "lme4" and "lmeTest"), as each participant viewed and rated four ads on a Likert scale. We ran a separate model for each independent variable as the combination of messenger and content type was not evenly distributed and some messenger/content combinations were not represented. Messenger type and content type were set as fixed effects and participant as a random effect; no covariates were included in the model. "Peer” and “Activation” were set as the reference categories for the messenger and content-type variables. A second set of models was run, including interaction terms, to assess whether messenger or content type effects varied by vaccination status.

## Results

### Demographics

A total of 113 participants participated in direct interviews and focus groups (Table 1). Most participants were Black or African American, married or living with a partner, and held a high school or bachelor’s degree. The majority were pregnant at the time of the study (61.9 percent) and were vaccinated for COVID-19 (76.3 percent).

**Table 1.** Demographic Characteristics of Participants

| Characteristic | Categories | N (%) |
| --- | --- | --- |
| Race and Ethnicity*<br>(N=92**) | Black or African American<br>Afro-Latinx<br>Mixed race/ethnicity with Black<br>or African American ethnicity | 62 (67.4)<br>14 (15.2)<br>7 (7.6) |
| Pregnancy Status (N=97) | Currently Pregnant<br>Pregnant within Last Six Months | 60 (61.9)<br>37 (38.1) |
| Marital Status (N=97) | Married<br>Not Married, Living with Partner<br>Not Married, not living with<br>partner<br>Single<br>Other | 37 (38.1)<br>21 (21.6)<br>5 (5.2)<br>31 (32)<br>3 (3.1) |
| Level of Education<br>(N=97) | Some High School<br>High School<br>Bachelor's Degree<br>Master's Degree<br>Trade School<br>Prefer not to Say | 12 (12.4)<br>32 (33)<br>40 (41.2)<br>8 (8.2)<br>1 (1)<br>4 (4.1) |
| Employment Status<br>(N=94) | Employed Full-Time<br>Employed Part-Time<br>Seeking Opportunities<br>Other<br>Prefer not to say | 23 (24.5)<br>30 (31.9)<br>22 (23.4)<br>12 (12.8)<br>5 (5.3) |
| Annual Household<br>Income (N=96) | < 25,000<br>25,000-50,000<br>50,000-100,000<br>100,000-200,000<br>Prefer not to say | 28 (29.2)<br>31 (32.3)<br>20 (20.8)<br>4 (4.2)<br>13 (13.5) |
| Religion (N=107) | Not Religious<br>Christian (Protestant)<br>Christian (Catholic)<br>Christian (Any other<br>denomination)<br>Judaism<br>Islam<br>Muslim<br>Other<br>Prefer not to say | 22 (20.6)<br>28 (26.2)<br>23 (21.5)<br>21 (19.6)<br>1 (0.9)<br>1 (0.9)<br>3 (2.8)<br>3 (2.8)<br>5 (4.7) |
| Political Affiliation (N=91) | Very Liberal | 12 (13.2) |
|  | Slightly Liberal<br>Slightly Conservative<br>Very Conservative<br>Prefer not to Say | 19 (20.9)<br>14 (15.4)<br>8 (8.8)<br>38 (41.8) |
| Vaccination Status<br>(N=97) | Has Received a COVID-19<br>Vaccine<br>Not Vaccinated | 74 (76.3)<br>23 (23.7) |
| Type of COVID-19<br>Vaccine Received<br>(N=68)** | Johnson & Johnson<br>Moderna<br>Pfizer<br>Other | 21 (30.9)<br>17 (23)<br>35 (51.5)<br>9 (13.2) |
| Number of Boosters<br>Received | 1<br>2<br>3<br>4 | 12 (17.6)<br>5 (7.4)<br>5 (7.4)<br>2 (2.9) |

\* Respondents were able to select more than one race or ethnicity
\*\* Respondents were able to select more than one type of vaccine received

### Main effects and interactions

The first set of models found no significant main effects for ad messenger or content-type as overall ratings across these dimensions were similar (Figure 2, Table 2, Models 1 and 2). Average ratings by messenger were most favorable for elder [mean (M)=2.08, SD=1.36] and least favorable for peer (M=2.24, SD=1.35). For content, social proof ads were most favorable (M=2.03, SD=1.30) and negative outcome ads were least favorable (M=2.32, SD=1.46). However, there was a large main effect of vaccination status, with non-vaccinated participants rating the ads less favorably (M=3.35, SD=1.52) than vaccinated participants (M=1.80, SD=1.02), indicating a lower likelihood of vaccination after seeing the ad. In a second set of models, we investigated interactions between vaccination status and ad messenger or ad content type. There was one significant interaction in which unvaccinated participants gave a more favorable rating for faith-based ads in comparison with peer messenger ads (M=3.07 vs. M=3.71, difference = -0.64), whereas vaccinated participants showed little difference between faith-based and peer messenger ads (M=1.86 vs. M=1.77, difference = +0.09; Figure 2, Table 3, p = 0.008). No other interaction terms were significant for messenger or content type. Overall, varying the ad messenger and content type did not lead to differences in the participants ratings of their likelihood to become vaccinated after seeing the ad.

**Figure 2.**
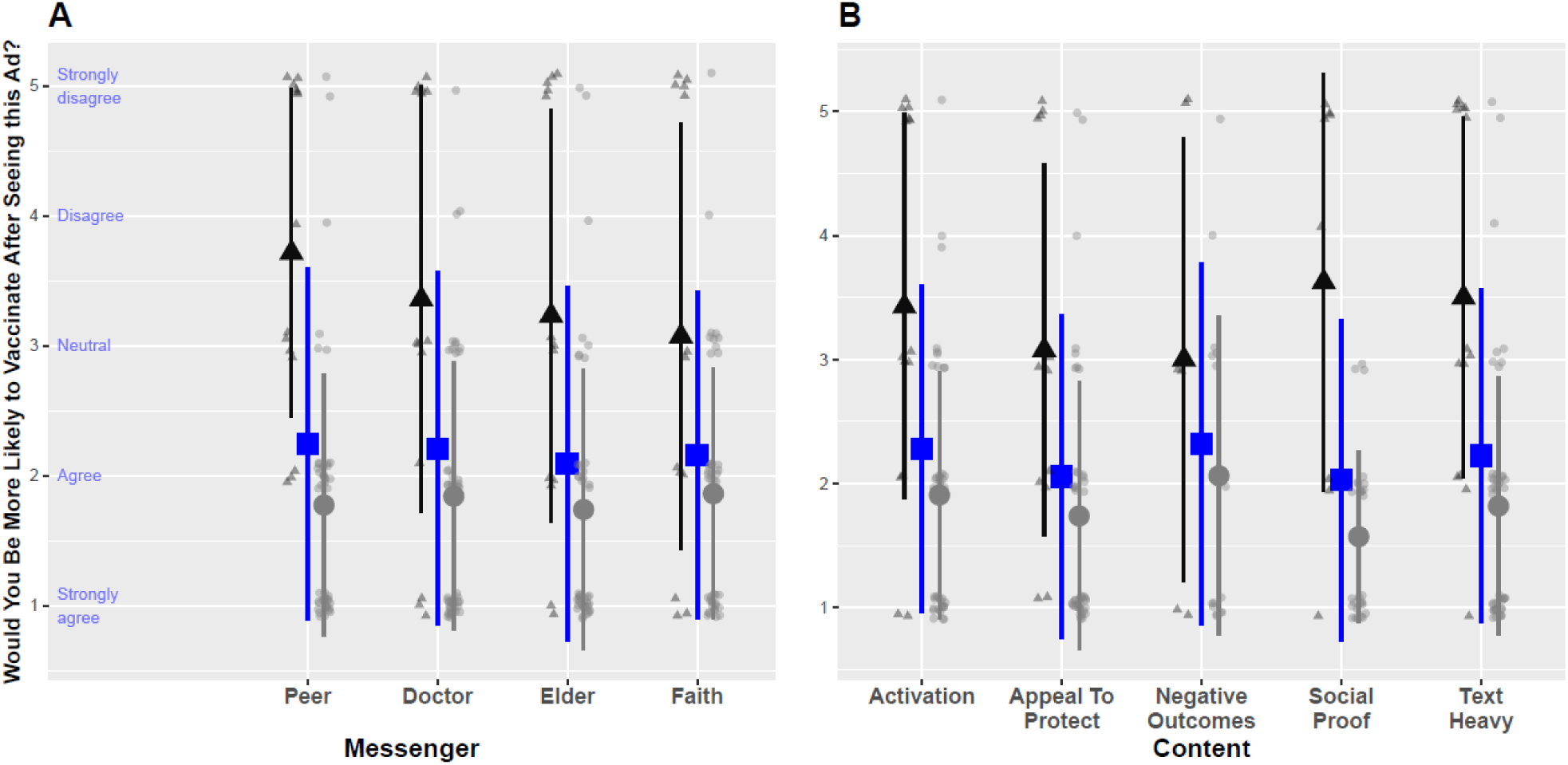
Subject Reported Likelihood of Becoming Vaccinated or Boosted After Seeing an Ad Featuring Specific Messengers and Content by Vaccination Status. This figure depicts the self-rated likelihood that a participant would receive a COVID-19 vaccine after seeing an ad showing a particular messenger (A) or a particular content type (B). The large blue square indicates the overall mean value in each category, black triangles and gray circles indicate group means for unvaccinated and vaccinated participants, respectively. The “negative outcomes” category reflected fear-based content. The Likert scale was constructed so that a rating of one indicated strong agreement that they were likely to become vaccinated after seeing the ad and a five rating that they strongly disagreed that they were likely to become vaccinated.

**Table 2.** Effect of Messenger Type and Content Type on Ad Ratings

| **Model 1: Ad Messenger Type and Vaccine Status** |  |  |  |  |  |
| --- | --- | --- | --- | --- | --- |
|  | How Likely to Vaccinate After Seeing the Ad |  |  |  |  |
| Predictors | Estimates | Std. Error | CI | Statistic | p |
| (Intercept) | 3.43 | 0.29 | 2.87 – 4.00 | 12.05 | **<0.001\*** |
| messenger [doctor] | -0.04 | 0.12 | -0.27 – 0.18 | -0.38 | 0.705 |
| messenger [elder] | -0.11 | 0.12 | -0.34 – 0.13 | -0.90 | 0.370 |
| messenger [faith] | -0.11 | 0.12 | -0.34 – 0.12 | -0.93 | 0.356 |
| vaccine status<br>[COVID19 vax? YES] | -1.55 | 0.32 | -2.17 – -<br>0.93 | -4.90 | **<0.001\*** |
| **Model 2: Ad Content Type and Vaccine Status** |  |  |  |  |  |
| Predictors | Estimates | Std. Error | CI | Statistic | p |
| (Intercept) | 3.47 | 0.29 | 2.91 – 4.04 | 12.14 | **<0.001\*** |
| content [social proof] | -0.09 | 0.14 | -0.35 – 0.18 | -0.65 | 0.517 |
| content [text heavy] | -0.07 | 0.12 | -0.30 – 0.15 | -0.65 | 0.518 |
| content<br>[appeal to protect] | -0.18 | 0.12 | -0.41 – 0.05 | -1.53 | 0.127 |
| content [negative outcome] | -0.25 | 0.17 | -0.58 – 0.07 | -1.52 | 0.129 |
| vaccine status<br>[COVID19 vax? YES] | -1.55 | 0.32 | -2.18 – -<br>0.92 | -4.88 | **<0.001\*** |

This table illustrates the interactions between vaccination status and participants' rating of their likelihood to become vaccinated after seeing the ad. Ratings corresponded as follows: a rating of 1 indicated "strongly agree", 2 "agree", 3 "neutral", 4 "disagree", and 5 "strongly disagree". In this model, the intercept is based on the unvaccinated group for either the peer (Model 1) and activation content (Model 2). There was a main effect for vaccine status, as vaccinated participants had significantly lower ratings, indicating a greater likelihood of vaccination after seeing the ad. \* $p < 0.05$ .

**Table 3.** Effect of Messenger Type by Vaccination Status on Ad Ratings

| Predictors | How Likely to Vaccinate After Seeing the Ad |  |  |  |  |
| --- | --- | --- | --- | --- | --- |
|  | Estimates | Std. Error | CI | Statistic | p-value |
| (Intercept) | 3.71 | 0.31 | 3.10 – 4.33 | 11.98 | <0.001* |
| messenger [doctor] | -0.36 | 0.23 | -0.81 – 0.10 | -1.55 | 0.123 |
| messenger [elder] | -0.37 | 0.24 | -0.84 – 0.09 | -1.58 | 0.116 |
| messenger [faith] | -0.64 | 0.23 | -1.10 – -0.19 | -2.78 | 0.006* |
| vaccine status yes/no<br>[COVID19 vax? YES] | -1.92 | 0.36 | -2.62 – -1.22 | -5.40 | <0.001* |
| messenger [doctor] ×<br>vaccine status yes/no<br>[COVID19 vax? YES] | 0.41 | 0.27 | -0.11 – 0.94 | 1.56 | 0.120 |
| messenger [elder] ×<br>vaccine status yes/no<br>[COVID19 vax? YES] | 0.35 | 0.27 | -0.18 – 0.89 | 1.30 | 0.194 |
| messenger [faith] ×<br>vaccine status yes/no<br>[COVID19 vax? YES] | 0.71 | 0.27 | 0.19 – 1.23 | 2.67 | 0.008* |

This table illustrates the interactions between vaccination status and participants' ratings of their likelihood to become vaccinated after seeing the ad. Ratings corresponded as follows: a rating of 1 indicated "strongly agree", 2 "agree", 3 "neutral", 4 "disagree", and 5 "strongly disagree". In this model, the intercept was the unvaccinated subjects when rating ads featuring peer messengers. Only the faith-based messenger was associated with a significantly greater likelihood of vaccination after seeing the ad, an effect driven by participants' vaccine status.
\*p<0.05.

There was, however, a significant interaction between participants’ vaccination status and the faith-based messenger (Table 3). Participants who were unvaccinated rated faith-based messengers more favorably than peer messengers, whereas vaccinated participants rated all messengers similarly and more favorably than unvaccinated participants. One participant said, “It makes me feel I should get vaccinated because it is coming from my pastor, and I have faith it would be effective.” She also reflected: “The ads have made me understand that different people have different ways of telling us what to do.” A different participant, also unvaccinated, responded to a faith-based activation ad by agreeing that “My pastor is someone I can trust.” Personal testimonials from faith leaders were also powerful drivers of vaccine uptake or hesitance. For example, one person stated: “I was sitting in church, and my pastor shared that he been vaccinated. And I say, ‘Well, God, that’s my sign.’ So, I was vaccinated.” In contrast, there was no interaction between vaccinated participants and messenger or ad content preference.

Overall, unvaccinated participants gave ads a higher average score than vaccinated participants, suggesting less favorable impressions of the ads. Most unvaccinated participants gave every ad that they saw the same rating, saying that they disagreed or strongly disagreed that they would be more likely to get vaccinated during pregnancy after seeing the ad. For example, one participant gave each of the four ads she saw a rating of 5, indicating that she strongly disagreed that it would encourage her to get vaccinated. For a social proof-content and peer messenger ad stating that hundreds of thousands of pregnant women have been safely vaccinated, this respondent said: “That’s great to know, but I’m a firm believer each person is different, and their bodies react differently.” In a text-heavy content ad with a care provider messenger, the same participant reiterated her earlier point: “I don’t agree with this information since everyone is different.” In response to an ‘appeal to protect ad’ with an elder messenger, she said: “I’ve taken other means to protect loved ones that [have] worked.” It was clear that differences in the messenger and the content did not change her overall opinion or distrust of the COVID-19 vaccine. Similarly, most vaccinated participants rated all of four ads that they saw equally favorably, regardless of changes in messengers or content.

### Qualitative Analysis

Analysis of interviews and focus groups identified three key themes with associated sub-themes that were influential in a participant’s decision to vaccinate during pregnancy: trust/mistrust, influence of health care providers, and medical racism (Table S4). Trusting one’s own opinions was a common answer among participants regarding the source they turned to for health information during pregnancy. Respondents noted receiving conflicting information and opinions from different sources and described sorting through this confusion by prioritizing themselves in decision-making. One participant described feeling overwhelmed by countless opinions and turning to herself, saying: “Every doctor in every clinic [has] their own diverse opinions about stuff, get their information from different places, so I just go with what I think is best for me.” Others recounted similarly individualized experiences saying, “I trust myself,” and, “You just think about what is good for you,” in response to questions about how they make health decisions in pregnancy.

Notably missing from many participants’ lists of trusted sources of information were physicians. When asked directly if physicians were a trusted source of information, many participants said that they were not. Though others did describe a trusting relationship with their OB or primary care physician, very few listed a physician as a trusted source of information. Interestingly, doulas were often included as trusted sources for a few participants, even those who had a strong distrust of doctors. Overall, however, the theme of trust was notable for a reliance on self and close relationships, and a mistrust or even fear of medical providers.

Another common theme in discussions of vaccination was the influence of medical racism. Participants described a general unease and mistrust of physicians, researchers, and novel medical advances like vaccines. They cited a history of medical racism as an influence for this unease and mistrust. One participant described her community’s fear of the vaccine: “What I’ve heard from families and friends is that it’s not for Black people. We shouldn’t take it because we don’t know what’s being put in it. I’ve heard things from about, you know, back in the day, experiments on Black people, right?” This participant specifically noted that a history of experimentation on Black people led to her, her family’s, and her friends’ hesitation to trust a vaccine that they do not have a lot of information about. Another participant highlighted a similar fear of being experimented on, saying, “I don’t trust the government. If we have to pay for a circumcision or ambulance and stuff, why is COVID and flu shots free all year round? They don’t care about us.” The fact that the vaccine was offered at no cost made her suspicious of the motivations of those providing the vaccine, and what those institutions might be gaining from vaccination.

While participants tended to rely on themselves for healthcare decisions, such as vaccination, a few individuals shared firsthand experiences that were crucial influences on their decision-making. A few participants said that the only reason she decided to get vaccinated was because of their pastor: “When my pastor unveiled that he took the vaccine, I took the vaccine.” It wasn’t the influence of any statistic or physician recommending the vaccine that led to uptake, but rather a trusted member of her community publicly revealing that he had personally decided to get vaccinated and that he was safe afterwards.

## Discussion

Our study indicates that unvaccinated participants were extremely likely to dislike all ad messengers and content types. In contrast, the vaccinated participants were highly likely to rate all ad messengers and content types as favorable in influencing them to receive the COVID-19 vaccination. Overall, the difference in ad ratings between vaccinated and unvaccinated participants was highly significant. However, the minimal variance in Likert score rating of the social media ads suggests that in the context of our study, participants were highly likely to rate the impact of social media ads on their likelihood of COVID-19 vaccination with a similar ad score; this data suggests that ad messenger and content type did not significantly affect participants’ ad ratings.

Vaccine hesitancy within Black communities has been observed in many studies.[9,19–21] This pattern of vaccine hesitancy within Black communities persisted with the COVID-19 vaccine. In a 2021 study, about 1 year after the release of the COVID-19 vaccine, a pooled review of vaccine hesitancy including more than 100,000 individuals in the United States demonstrated that vaccine hesitancy overall in the United States was 26.3% and notably higher among Black (41.6%) and Hispanic (30.2%) individuals.[22] COVID-19 vaccine hesitancy is even more pronounced among pregnant individuals, with studies reporting COVID-19 vaccine acceptance rates of only 44.3-58.35%, compared to 76.2% in non-pregnant individuals.[23–25] Black race was associated with hesitancy towards the vaccine during pregnancy.[25] While our study did not specifically evaluate vaccine uptake rates in our study population, our qualitative results support the findings of vaccine hesitancy. The idea that there is not one universal determinant for vaccine hesitancy in Black communities is evident in other articles that qualitatively explored reasons for vaccine hesitancy and mistrust of the vaccine.[9,21,26] As supported by our own interviews, vaccine hesitancy was influenced by a wide variety of factors including, but not limited to, mistrust of the medical industry, medical racism, myths about the vaccine and concern about its safety, and fears about the speed of the vaccine development.

Among Black pregnant and postpartum women, many factors were cited as driving their COVID-19 vaccine hesitancy, including individual-person beliefs influenced by ethnicity, culture, family, and religion, and vaccine disinformation.[9,22] It is possible that our social media advertisements failed to influence vaccine-hesitant individuals because the reasons underlying their hesitancy were diverse. The advertisements may not have addressed the individuals’ complex concerns. A prior study indicated that using messengers from the Black community, such as Black faith leaders and Black physicians, were effective in promoting vaccination.[27] Although race concordance between physicians and patients has been shown across studies to be associated with greater patient satisfaction and perception of care, the Black messengers in our study were not trusted by the unvaccinated participants.[28,29] Obstetrical care is one of great sensitivity for many patients, and reviews of perinatal and obstetric experiences for Black women have been largely negative with experiences of racism and biases in care.[30–32]

As many participants referenced their trust and support of their doula, it is possible that among pregnant Black women, a doula may be a better medical professional messenger for vaccine promotion.[35,36] Our study did not specifically address the use of a doula as a messenger of public health information. Overall, given that individuals offered multifactorial reasons for vaccine hesitancy, each influenced by the individual’s own healthcare experience and race. Therefore, there may not be a “one size fits all” approach to public health messaging in marginalized communities that have experienced racism and mistrust of healthcare providers. Our study also did not identify a variable, such as the messenger or the message tone, that could be altered to make a social media advertisement more influential. The key to successful messaging, in turn, may lie with emphasizing the perspectives, cultural and faith-related values of individuals directly from these communities to minimize the potential for the influence of medical racism in decision-making. As unvaccinated participants preferred the faith leader messenger, faith-based organizations may have considerable impact on vaccine response in a public health emergency.[37,38]

A strength of this study is the focus on the opinions and reactions of Black pregnant individuals to vaccine messaging, who are underrepresented in scientific studies and have the lowest vaccination rates in pregnancy. Another strength of our study was the use of Black community health advocates and partners to conduct focus groups and interviews. These individuals and organizations were chosen as they were trusted within the Black community and likely to elicit honest responses to the interview questions. However, many of these individuals had backgrounds in community health rather than in research, and therefore follow-up questions to elicit more in-depth responses to some questions were not always posed. Further, the interviews were often performed in batches and quality could not be controlled in real time. Sometimes, it was difficult to discern exactly how many participants’ voices were captured during the interviews. As a result, there was a mismatch between the number of REDCap surveys (quantitative results for ad scores) and the number of participants in qualitative interviews/focus groups. Despite these limitations, we believe that strong community partnerships and trust are essential components to performing a qualitative study in an underserved and vaccine hesitant population. An unexpected finding and study limitation was that the quantitative results had minimal inter-subject and group variability with COVID-19 vaccinated participants viewing ads more favorably and unvaccinated participants perceiving ads negatively. Polarization of opinion by vaccination status made it difficult to use the quantitative results to determine which ad messenger or content type was most influential. Future studies will need to anticipate these issues in the study design to enhance success of understanding factors influencing vaccine hesitancy in Black pregnant individuals to build an effective public health messaging campaign.

Involving trusted individuals in the Black community from faith-based organizations may be one step toward improving trust within the medical community and reducing vaccine hesitancy. Many studies have demonstrated the potential to work with community leaders, such as church pastors in faith-based communities, to increase COVID-19 vaccine equity and share public health information.[27,43] Future research could evaluate the efficacy of these interventions, as well as public health messaging that features a specific trusted community figure rather than a generic “faith leader” or “healthcare provider.” Many individuals, including those who were vaccine-hesitant, also mentioned their doula as a trusted source of information. Public health campaigns featuring doulas may engender greater trust in the Black pregnant community and might provide a unique and trusted avenue for vaccine promotion.

In summary, we were unable to identify a clearly effective messenger for COVID-19 vaccine promotion in Black pregnant people, as we have in prior studies.[16,17] Race-congruent physician messengers were not a source of trusted information in this community, as they were for rural Hispanic pregnant individuals in a previous study in the Western U.S. using similar methodology.[17] Similarly, peers were not a trusted source of information as they had been for White pregnant people living in the rural U.S.[16] The singular most crucial determinant of how participants viewed COVID-19 vaccination ads during pregnancy was their own vaccination status: vaccinated individuals viewed the ads positively, whereas unvaccinated individuals viewed them negatively. Although unvaccinated individuals rated ads less favorably than their vaccinated peers, faith-based messengers were rated more favorably by unvaccinated participants. Participants reported many factors contributing to their COVID-19 vaccine hesitancy, including mistrust of the medical system, concern about medical racism, belief in vaccine myths, concern about vaccine safety, and fears about the speed of vaccine development. Moving forward, greater efforts to connect with pastors and other community leaders to build trust in vaccination and other preventive care measures is critical before the next pandemic. Addressing common vaccine myths is also necessary, as it may help prevent vaccine disinformation from taking hold. Finally, a greater effort to understand how diverse pregnant populations perceive vaccine information is essential to meet the higher threshold for vaccine acceptance in pregnancy.[5] Continued efforts to build trust with Black pregnant people at every level is crucial to improving their mortality and preventing needless deaths due to vaccine hesitancy.

## Acknowledgments

We are incredibly grateful to the Black pregnant and recently pregnant individuals who participated in our study, shared their individual experiences, and offered their thoughts and views on vaccination in pregnancy. Without the strong partnership from Black Health, Inc., this work would not have been possible. Within Black Health, we acknowledge and thank a team of individuals for their efforts in supporting the study: Evelyn Botwe, Rachel Johnson, Alexandra Quintero, Dinah Aryeh and Disleiry Benitez. We also thank the dedicated individuals who assisted with the study at the Afiya Center (Dallas, TX), Mothering Justice (Detroit, MI), New York Police Department Community Engagement Office (New York, NY).

## Author Contributions

Conceptualization, E.C., M.S., J.M., R.J., K.A.W.; methodology, E.C., E.E., M.S., J.M., S.S., A.S., R.J., K.A.W.; formal analysis, E.C., C.B., I.C., E.E., J.M., K.A.W.; investigation, E.C., E.E., R.J., M.B., M.S., C.B., I.C., S.S., J.M., A.S., K.A.W.; data curation, E.C., E.E., R.J., J.M.; writing—original draft preparation, E.C., E.E., C.B., I.C., K.A.W.; writing—review and editing, E.C., E.E., R.J., M.B., M.S., C.B., I.C., S.S., J.M., A.S., K.A.W.; supervision, E.C., M.B., R.J., K.A.W.; project administration, K.A.W., M.B., R.J.; funding acquisition, K.A.W. All authors have read and agreed to the published version of the manuscript.

## Institutional Review Board Statement

The study was conducted in accordance with the Declaration of Helsinki and approved by the Institutional Review Board of the University of Washington (STUDY00014356, initial approval 11/05/2021).

## Informed Consent Statement

We received informed consent from all respondents to participate in the study before starting the interviews.

## Data Availability Statement

The data presented in this study are available on request from the corresponding author. The audio interviews themselves are not available due to privacy.

## REFERENCES

1. Siston, A.M.; Rasmussen, S.A.; Honein, M.A.; Fry, A.M.; Seib, K.; Callaghan, W.M.; Louie, J.; Doyle, T.J.; Crockett, M.; Lynfield, R.;, et al. Pandemic 2009 influenza A(H1N1) virus illness among pregnant women in the United States. JAMA : the journal of the American Medical Association 2010, *303*, 1517–1525, doi:10.1001/jama.2010.479.

2. Ribeiro, A.F.; Pellini, A.C.G.; Kitagawa, B.Y.; Marques, D.; Madalosso, G.; Fred, J.; Albernaz, R.K.M.; Carvalhanas, T.; Zanetta, D.M.T. Severe influenza A(H1N1)pdm09 in pregnant women and neonatal outcomes, State of Sao Paulo, Brazil, 2009. PloS one 2018, 13, e0194392, doi:10.1371/journal.pone.0194392.

3. Jamieson, D.J.; Honein, M.A.; Rasmussen, S.A.; Williams, J.L.; Swerdlow, D.L.; Biggerstaff, M.S.; Lindstrom, S.; Louie, J.K.; Christ, C.M.; Bohm, S.R.;, et al. H1N1 2009 influenza virus infection during pregnancy in the USA. Lancet 2009, 374, 451–458, doi:10.1016/S0140-6736(09)61304-0.

4. Gunnes, N.; Gjessing, H.K.; Bakken, I.J.; Ghaderi, S.; Gran, J.M.; Hungnes, O.; Magnus, P.; Samuelsen, S.O.; Skrondal, A.; Stoltenberg, C.;, et al. Seasonal and pandemic influenza during pregnancy and risk of fetal death: A Norwegian registry-based cohort study. Eur J Epidemiol 2020, 35, 371–379, doi:10.1007/s10654-020-00600-z.

5. Cox, E.; Sanchez, M.; Taylor, K.; Baxter, C.; Crary, I.; Every, E.; Futa, B.; Adams Waldorf, K.M. A Mother’s Dilemma: The 5-P Model for Vaccine Decision-Making in Pregnancy. Vaccines (Basel) 2023, 11, doi:10.3390/vaccines11071248.

6. Alcendor, D.J.; Matthews-Juarez, P.; Smoot, D.; Hildreth, J.E.K.; Tabatabai, M.; Wilus, D.; Brown, K.Y.; Juarez, P.D. The COVID-19 Vaccine and Pregnant Minority Women in the US: Implications for Improving Vaccine Confidence and Uptake. Vaccines (Basel) 2022, 10, doi:10.3390/vaccines10122122.

7. Reifferscheid, L.; Marfo, E.; Assi, A.; Dube, E.; MacDonald, N.E.; Meyer, S.B.; Bettinger, J.A.; Driedger, S.M.; Robinson, J.; Sadarangani, M.;, et al. COVID-19 vaccine uptake and intention during pregnancy in Canada. Can J Public Health 2022, 113, 547–558, doi:10.17269/s41997-022-00641-9.

8. Battarbee, A.N.; Stockwell, M.S.; Varner, M.; Newes-Adeyi, G.; Daugherty, M.; Gyamfi-Bannerman, C.; Tita, A.T.; Vorwaller, K.; Vargas, C.; Subramaniam, A.;, et al. Attitudes Toward COVID-19 Illness and COVID-19 Vaccination among Pregnant Women: A Cross-Sectional Multicenter Study during August-December 2020. American journal of perinatology 2022, 39, 75–83, doi:10.1055/s-0041-1735878.

9. Avorgbedor, F.; Gondwe, K.W.; Aljarrah, A.; Bankole, A.O. COVID-19 Vaccine Decision-Making Among Black Pregnant and Postpartum Women. J Racial Ethn Health Disparities 2023, doi:10.1007/s40615-023-01675-6.

10. Marcell, L.; Dokania, E.; Navia, I.; Baxter, C.; Crary, I.; Rutz, S.; Soto Monteverde, M.J.; Simlai, S.; Hernandez, C.; Huebner, E.M., et al. One Vax Two Lives: A Social Media Campaign and Research Program to Address COVID-19 Vaccine Hesitancy in Pregnancy. American journal of obstetrics and gynecology 2022, doi:10.1016/j.ajog.2022.06.022.

11. Gaskin, D.J.; Dinwiddie, G.Y.; Chan, K.S.; McCleary, R.R. Residential segregation and the availability of primary care physicians. Health Serv Res 2012, 47, 2353–2376, doi:10.1111/j.1475-6773.2012.01417.x.

12. Cohen, R.A.; Terlizzi, E.P. Demographic Variation in Health Insurance Coverage:United States, 2022. Natl Health Stat Report 2023, 1–15.

13. Scharff, D.P.; Mathews, K.J.; Jackson, P.; Hoffsuemmer, J.; Martin, E.; Edwards, D. More than Tuskegee: understanding mistrust about research participation. J Health Care Poor Underserved 2010, 21, 879–897, doi:10.1353/hpu.0.0323.

14. Dawson, M.A.; Giger, J.N.; Powell-Young, Y.; Brannon, C.B. Why African-Americans are Hesitant to Take the Newly Proposed COVID-19 Vaccines: Tuskegee Revisited. J Natl Black Nurses Assoc 2020, 31, vi-viii.

15. Alsan, M.; Wanamaker, M.; Hardeman, R.R. The Tuskegee Study of Untreated Syphilis: A Case Study in Peripheral Trauma with Implications for Health Professionals. Journal of general internal medicine 2020, 35, 322–325, doi:10.1007/s11606-019-05309-8.

16. Cox, E.; Sanchez, M.; Baxter, C.; Crary, I.; Every, E.; Munson, J.; Stapley, S.; Stonehill, A.; Taylor, K.; Widmann, W., et al. COVID-19 Vaccine Hesitancy among English-Speaking Pregnant Women Living in Rural Western United States. Vaccines (Basel) 2023, 11, doi:10.3390/vaccines11061108.

17. Sanchez, M.; Martel, I.; Cox, E.; Crary, I.; Baxter, C.; Every, E.; Munson, J.; Stapley, S.; Stonehill, A.; Adams Waldorf, K.M. Factors Influencing COVID-19 Vaccine Uptake among Spanish-Speaking Pregnant People. Vaccines (Basel) 2023, 11, doi:10.3390/vaccines11111726.

18. Saha, S.; Beach, M.C. Impact of Physician Race on Patient Decision-Making and Ratings of Physicians: a Randomized Experiment Using Video Vignettes. Journal of general internal medicine 2020, 35, 1084–1091, doi:10.1007/s11606-020-05646-z.

19. Cenat, J.M.; Noorishad, P.G.; Bakombo, S.M.; Onesi, O.; Mesbahi, A.; Darius, W.P.; Caulley, L.; Yaya, S.; Chomienne, M.H.; Etowa, J.;, et al. A Systematic Review on Vaccine Hesitancy in Black Communities in Canada: Critical Issues and Research Failures. Vaccines (Basel) 2022, 10, doi:10.3390/vaccines10111937.

20. Willis, D.E.; Andersen, J.A.; Montgomery, B.E.E.; Selig, J.P.; Shah, S.K.; Zaller, N.; Bryant-Moore, K.; Scott, A.J.; Williams, M.; McElfish, P.A. COVID-19 Vaccine Hesitancy and Experiences of Discrimination Among Black Adults. J Racial Ethn Health Disparities 2023, 10, 1025–1034, doi:10.1007/s40615-022-01290-x.

21. Sekimitsu, S.; Simon, J.; Lindsley, M.M.; Jones, M.; Jalloh, U.; Mabogunje, T.; Kerr, J.; Willingham, M.; Ndousse-Fetter, S.B.; White-Hammond, G., et al. Exploring COVID-19 Vaccine Hesitancy Amongst Black Americans: Contributing Factors and Motivators. Am J Health Promot 2022, 36, 1304–1315, doi:10.1177/08901171221099270.

22. Khubchandani, J.; Macias, Y. COVID-19 vaccination hesitancy in Hispanics and African-Americans: A review and recommendations for practice. Brain Behav Immun Health 2021, 15, 100277, doi:10.1016/j.bbih.2021.100277.

23. Yasmin, F.; Najeeb, H.; Moeed, A.; Naeem, U.; Asghar, M.S.; Chughtai, N.U.; Yousaf, Z.; Seboka, B.T.; Ullah, I.; Lin, C.Y., et al. COVID-19 Vaccine Hesitancy in the United States: A Systematic Review. Front Public Health 2021, 9, 770985, doi:10.3389/fpubh.2021.770985.

24. Sutton, D.; D’Alton, M.; Zhang, Y.; Kahe, K.; Cepin, A.; Goffman, D.; Staniczenko, A.; Yates, H.; Burgansky, A.; Coletta, J., et al. COVID-19 vaccine acceptance among pregnant, breastfeeding, and nonpregnant reproductive-aged women. Am J Obstet Gynecol MFM 2021, 3, 100403, doi:10.1016/j.ajogmf.2021.100403.

25. Levy, A.T.; Singh, S.; Riley, L.E.; Prabhu, M. Acceptance of COVID-19 vaccination in pregnancy: a survey study. Am J Obstet Gynecol MFM 2021, 3, 100399, doi:10.1016/j.ajogmf.2021.100399.

26. Scales, D.; Gorman, S.; Windham, S.; Sandy, W.; Gregorian, N.; Hurth, L.; Radhakrishnan, M.; Akunne, A.; Gorman, J.M. ’They’ve all endorsed it…but I’m just not there:’ a qualitative exploration of COVID-19 vaccine hesitancy reported by Black and Latinx individuals. BMJ Open 2023, 13, e072619, doi:10.1136/bmjopen-2023-072619.

27. Dada, D.; Djiometio, J.N.; McFadden, S.M.; Demeke, J.; Vlahov, D.; Wilton, L.; Wang, M.; Nelson, L.E. Strategies That Promote Equity in COVID-19 Vaccine Uptake for Black Communities: a Review. J Urban Health 2022, 99, 15–27, doi:10.1007/s11524-021-00594-3.

28. Loeb, S.; Ravenell, J.E.; Gomez, S.L.; Borno, H.T.; Siu, K.; Sanchez Nolasco, T.; Byrne, N.; Wilson, G.; Griffith, D.M.; Crocker, R., et al. The Effect of Racial Concordance on Patient Trust in Online Videos About Prostate Cancer: A Randomized Clinical Trial. JAMA Netw Open 2023, 6, e2324395, doi:10.1001/jamanetworkopen.2023.24395.

29. Ku, L.; Vichare, A. The Association of Racial and Ethnic Concordance in Primary Care with Patient Satisfaction and Experience of Care. Journal of general internal medicine 2023, 38, 727–732, doi:10.1007/s11606-022-07695-y.

30. Chambers, B.D.; Fontenot, J.; McKenzie-Sampson, S.; Blebu, B.E.; Edwards, B.N.; Hutchings, N.; Karasek, D.; Coleman-Phox, K.; Curry, V.C.; Kuppermann, M. "It was just one moment that I felt like I was being judged": Pregnant and postpartum black Women’s experiences of personal and group-based racism during the COVID-19 pandemic. Soc Sci Med 2023, 322, 115813, doi:10.1016/j.socscimed.2023.115813.

31. OjiNjideka Hemphill, N.; Crooks, N.; Zhang, W.; Fitter, F.; Erbe, K.; Rutherford, J.N.; Liese, K.L.; Pearson, P.; Stewart, K.; Kessee, N.;, et al. Obstetric experiences of young black mothers: An intersectional perspective. Soc Sci Med 2023, 317, 115604, doi:10.1016/j.socscimed.2022.115604.

32. Nguyen, T.T.; Criss, S.; Kim, M.; De La Cruz, M.M.; Thai, N.; Merchant, J.S.; Hswen, Y.; Allen, A.M.; Gee, G.C.; Nguyen, Q.C. Racism During Pregnancy and Birthing: Experiences from Asian and Pacific Islander, Black, Latina, and Middle Eastern Women. J Racial Ethn Health Disparities 2023, 10, 3007–3017, doi:10.1007/s40615-022-01475-4.

33. Quinn, S.C.; Jamison, A.; Freimuth, V.S.; An, J.; Hancock, G.R.; Musa, D. Exploring racial influences on flu vaccine attitudes and behavior: Results of a national survey of White and African American adults. Vaccine 2017, 35, 1167–1174, doi:10.1016/j.vaccine.2016.12.046.

34. Rosenthal, L.; Lobel, M. Gendered racism and the sexual and reproductive health of Black and Latina Women. Ethn Health 2020, 25, 367–392, doi:10.1080/13557858.2018.1439896.

35. Rice, H.; Collins, C.; Cherney, E. Beyond Birth Work: Addressing Social Determinants of Health With Community Perinatal Support Doulas. Clin Nurs Res 2024, 10547738241244590, doi:10.1177/10547738241244590.

36. Louis-Jacques, A.F.; Applequist, J.; Perkins, M.; Williams, C.; Joglekar, R.; Powis, R.; Daniel, A.; Wilson, R. Florida Doulas’ Perspectives on Their Role in Reducing Maternal Morbidity and Health Disparities. Womens Health Issues 2024, doi:10.1016/j.whi.2024.01.003.

37. Syed, U.; Kapera, O.; Chandrasekhar, A.; Baylor, B.T.; Hassan, A.; Magalhaes, M.; Meidany, F.; Schenker, I.; Messiah, S.E.; Bhatti, A. The Role of Faith-Based Organizations in Improving Vaccination Confidence & Addressing Vaccination Disparities to Help Improve Vaccine Uptake: A Systematic Review. Vaccines (Basel) 2023, 11, doi:10.3390/vaccines11020449.

38. Chu, J.; Pink, S.L.; Willer, R. Religious identity cues increase vaccination intentions and trust in medical experts among American Christians. Proc Natl Acad Sci U S A 2021, 118, doi:10.1073/pnas.2106481118.

39. Goes, E.F.; Ferreira, A.J.F.; Ramos, D. Anti-Black racism and maternal death from COVID-19: what have we seen in the Pandemic? Cien Saude Colet 2023, 28, 2501–2510, doi:10.1590/1413-81232023289.08412022.

40. Simpson, K.R. Effect of the COVID-19 Pandemic on Maternal Health in the United States. MCN Am J Matern Child Nurs 2023, 48, 61, doi:10.1097/NMC.0000000000000895.

41. Piekos, S.N.; Hwang, Y.M.; Roper, R.T.; Sorensen, T.; Price, N.D.; Hood, L.; Hadlock, J.J. Effect of COVID-19 vaccination and booster on maternal-fetal outcomes: a retrospective cohort study. Lancet Digit Health 2023, 5, e594–e606, doi:10.1016/S2589-7500(23)00093-6.

42. Morgan, J.A.; Biggio, J.R., Jr.; Martin, J.K.; Mussarat, N.; Chawla, H.K.; Puri, P.; Williams, F.B. Maternal Outcomes After Severe Acute Respiratory Syndrome Coronavirus 2 (SARS-CoV-2) Infection in Vaccinated Compared With Unvaccinated Pregnant Patients. Obstet Gynecol 2022, 139, 107–109, doi:10.1097/AOG.0000000000004621.

43. Moore, D.; Mansfield, L.N.; Onsomu, E.O.; Caviness-Ashe, N. The Role of Black Pastors in Disseminating COVID-19 Vaccination Information to Black Communities in South Carolina. International journal of environmental research and public health 2022, 19, doi:10.3390/ijerph19158926.

44. Street, R.L., Jr.; O’Malley, K.J.; Cooper, L.A.; Haidet, P. Understanding concordance in patient-physician relationships: personal and ethnic dimensions of shared identity. Ann Fam Med 2008, 6, 198–205, doi:10.1370/afm.821.

