## Supplemental Information for "Understanding COVID-19 Vaccine Hesitancy Among Black and Afro-Latinx Pregnant Individuals: A Mixed-Methods Approach Utilizing Focus Groups and Social Media Ad Reaction"

**Table S1. Number of Subjects Viewing Specific Combinations of Ad Messenger and Ad Content**

| Messenger | Ad Number | Content | Number of Views |
|---|---|---|---|
| Peer | 1 | Appeal to Protect | 5 |
| | 2 | Text-Heavy | 5 |
| | 3 | Social Proof | 10 |
| | 4 | Information (Negative outcomes) | 5 |
| | 5 | Activation | 5 |
| Elder | 6 | Appeal to Protect | 13 |
| | 7 | Text-Heavy | 6 |
| | 8 | Activation | 9 |
| Doctor | 9 | Appeal to Protect | 7 |
| | 10 | Text-Heavy | 6 |
| | 11 | Social Proof | 9 |
| | 12 | Information (Negative outcomes) | 6 |
| | 13 | Activation | 2 |
| Faith* | 14 | Appeal to Protect | 4 |
| | 15 | Text-Heavy | 11 |
| | 16 | Activation | 14 |

*The faith ad was designed to match the participant's self-reported religion: Pastor (Protestant or Any Other Christian Faith), Priest (Catholic), and Imam (Muslim).

**Table S2. Focus Group Interview Guide**

| **Question Theme** | **Questions** |
|---|---|
| Pregnancy | Tell me about how this pregnancy/ postpartum is going.<br>Walk me through your biggest challenges right now. |
| COVID-19 Vaccine | Tell me what you have heard about the COVID-19 vaccine from family and friends.<br>Please explain why you did or didn't believe [what you have heard].<br>Tell me what you heard about the COVID-19 vaccine from a healthcare worker during your pregnancy.<br>Please explain why you did or didn't believe [what you have heard].<br>For those who are part of a Church or faith-based community, tell me what you have heard about the COVID-19 vaccine from members of your faith-based community? Did you believe it? Why or why not? |
| COVID-19 Experience | Has anyone known people who have had COVID-19?<br>Describe your experiences with the COVID-19 disease either for yourself or others.<br>How concerned are you (or were you) about catching COVID-19 during your pregnancy?<br>***If concerned***, what were you worried might have happened to you or the fetus? ***If not concerned***, tell me more about why you weren't worried. |
| Decision Making in Pregnancy | Tell me who you generally trust for health information and why.<br>Walk me through how you made the decision to vaccinate yourself or not vaccinate for COVID-19?<br>Follow-up questions: Was anyone's opinion important in helping you make your decision? Was there anything you read or saw that influenced your decision? Please tell me more.<br>**If unvaccinated, partially vaccinated or unboosted**, do you have plans to get vaccinated or boosted for COVID-19, complete your vaccine series or get boosted? |

**Table S3. Thematic Codebook**

| Parent Code | Description |
|---|---|
| Distrusted Sources of Health Information | Participant mentions distrusted sources of health information |
| Trusted Sources of Health Information | Participant mentions trusted sources of health information |
| Experiences with COVID-19 for Self or Others | Participant describes their own experiences having COVID-19, or observing someone (near them) else's experience with COVID |
| Self-perceived Risk of COVID-19 | Participant describes self-perceived risk of COVID-19 |
| Barriers to Vaccination | Participant describes perceived barriers to getting vaccinated |
| Facilitators/cues to action | Participant describes perceived facilitators to vaccination or cues to action |
| Vaccine Hesitant | Participant describes hesitancy of vaccines or sources of vaccine hesitancy |
| Vaccine Positive | Participant describes feeling positively toward vaccines |
| Interactions with health care provider | Participant describes experiences with health care provider |
| Experiences with COVID-19 vaccine | Participant describes experiences receiving the COVID vaccine, either for themselves or those around them |
| Challenges in Pregnancy | Participant describes challenges faced in their pregnancies |

**Table S4. Key Themes, Sub-Themes, and Quotes**

| Theme | Associated sub-theme | Supporting Quotes |
|---|---|---|
| Trust | Trust in Self<br>Trust in Family<br>Trust in Doulas | "Every doctor in every clinic [has] their own diverse opinions about stuff, get their information from different places, so I just go with what I think is best for me."<br>"I trust myself." |
| | | "My mom used to [tell] me all the time, 'you should get it. You got the opportunity to get it. Go get it." |
| | | In response to who is a trusted source of information: "myself, my doula, my family."<br>"If you notice most doulas are dark skinned, and the government don't like dark skinned people. So that's who you have to trust." |
| Mistrust | History of Medical Racism<br>Mistrust of Government<br>Mistrust of Healthcare system | "What I've heard from families and friends is that it's not for Black people. We shouldn't take it because we don't know what's being put in it. I've heard things from about, you know, back in the day experiments on Black people, right?" |
| | | "I don't trust the government. If we have to pay for a circumcision or ambulance and stuff, why is COVID and flu shots free all year round? They don't care about us." |
| Personal experience | Positive Influence<br>Negative Influence | "When my pastor unveiled that he took the vaccine, I took the vaccine." |
| | | "I've actually just went to the doctors, and he was telling us that pediatricians recommend you to get the vaccine. But he said he [the doctor] understands, because he said if his wife was pregnant, he wouldn't push it [the vaccine] or wouldn't want it right now." |

**Figure S1. Enlarged Social Media Ads with an Appeal to Protect Message.**

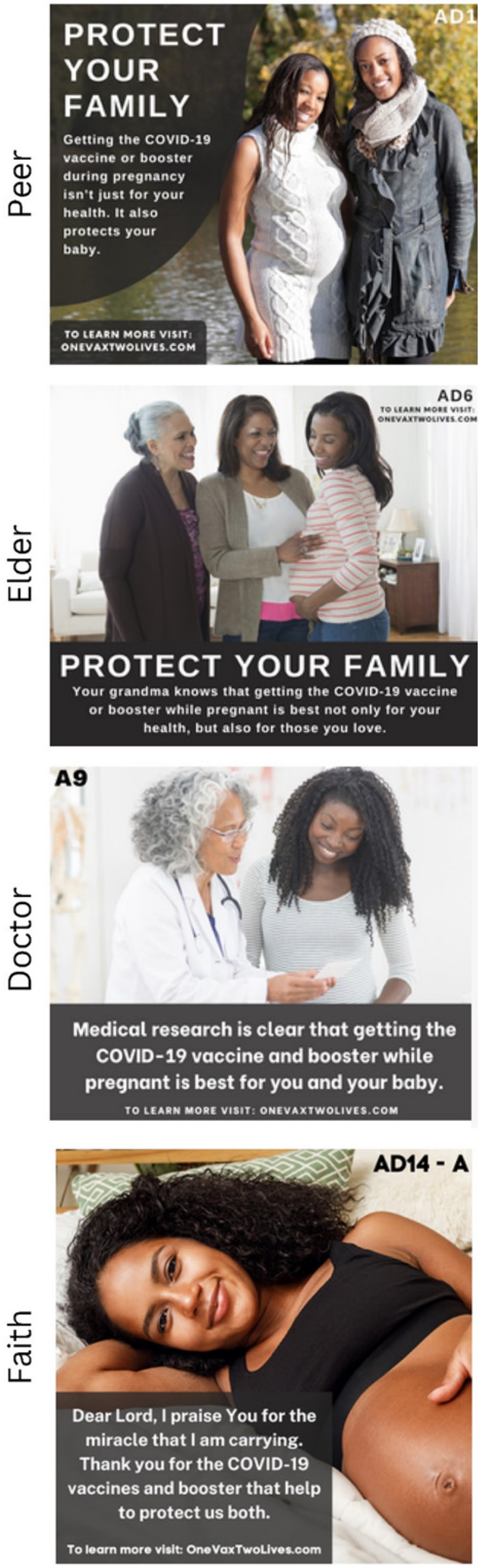

This figure depicts ads featuring different messengers that have appeal to the viewer to protect their family by vaccinating or boosting themselves with the COVID-19 vaccine in pregnancy.

**Figure S2. Enlarged Social Media Ads with a Text Heavy Message**

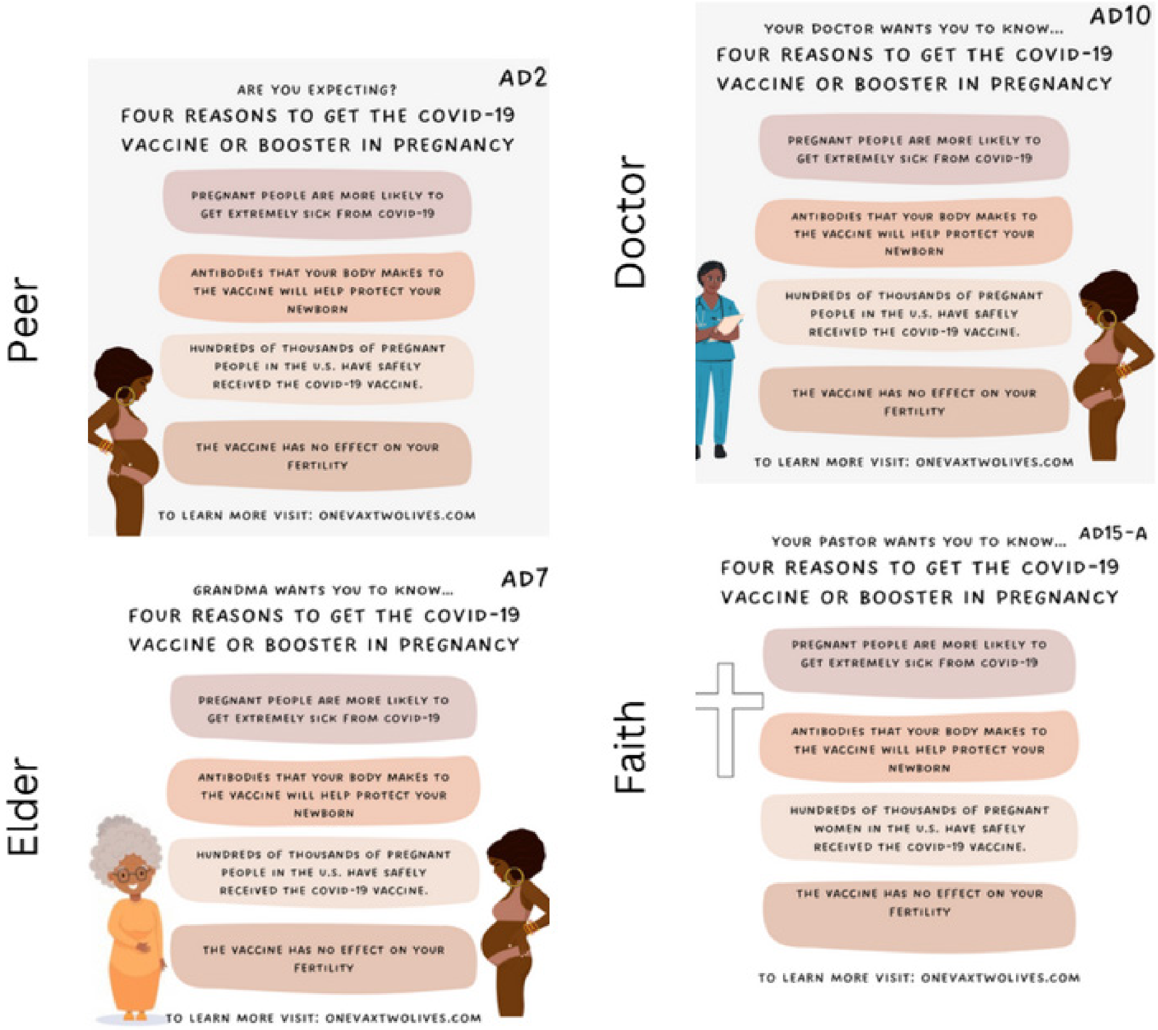

This figure depicts sample social media advertisements featuring a text heavy message promoting the COVID-19 vaccine in pregnancy in combination with different messengers.

**Figure S3. Enlarged Social Media Ads with a Social Proof Message**

Peer

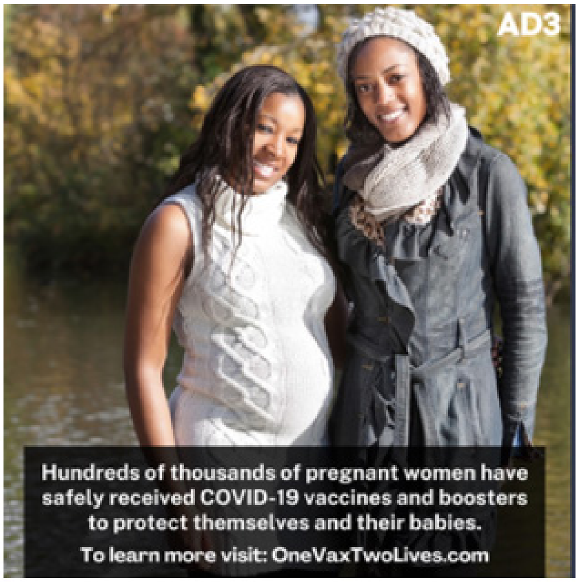

Doctor

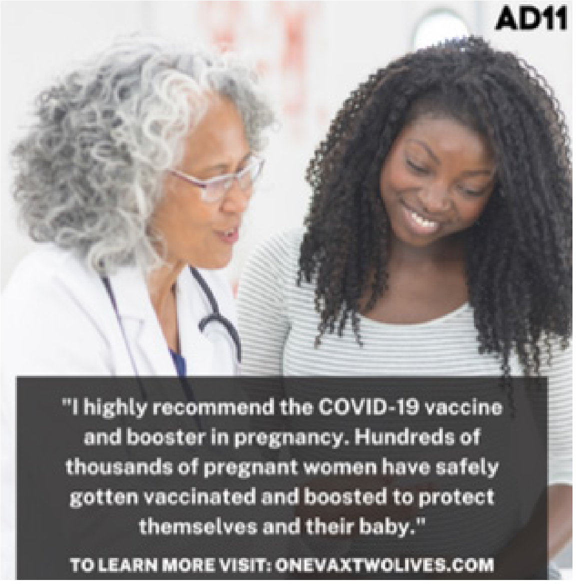

This figure depicts sample social media advertisements featuring a message emphasizing “social proof” by stating that hundreds of thousands of pregnant women had safely received the vaccine. Some messenger combinations were not realistic and were, therefore, not created or shown for this particular ad content type.

**Figure S4. Enlarged Social Media Ads with Information About Negative Outcomes**

Peer

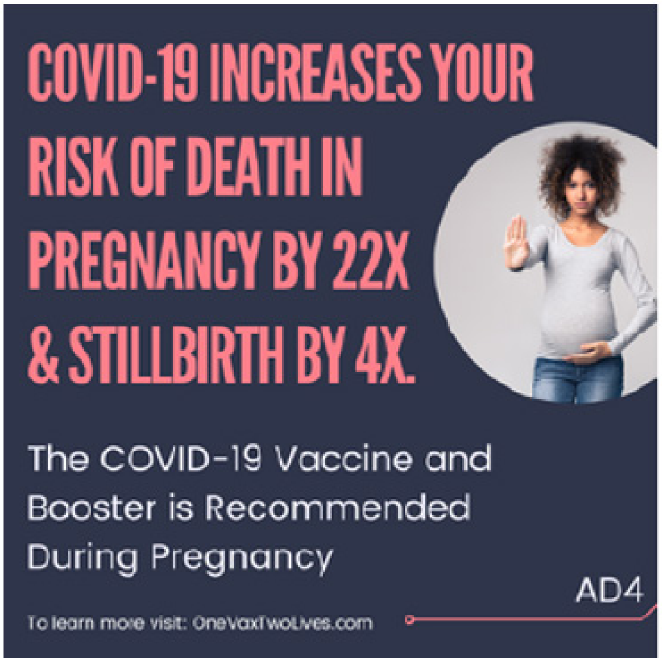

Doctor

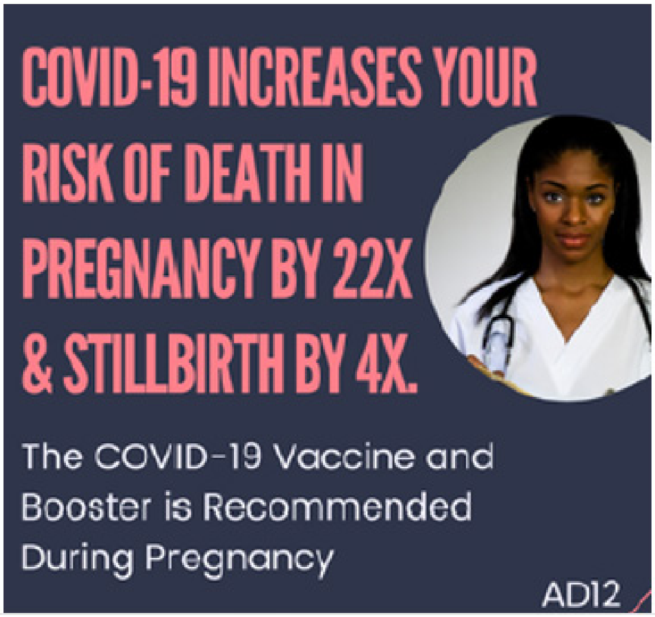

This figure depicts sample social media advertisements featuring messages emphasizing the negative outcomes that can occur if a pregnant person contracts COVID-19. Some messenger combinations were not realistic for this ad and were, therefore, not created.

**Figure S5. Enlarged Social Media Ads with an Activation Message**

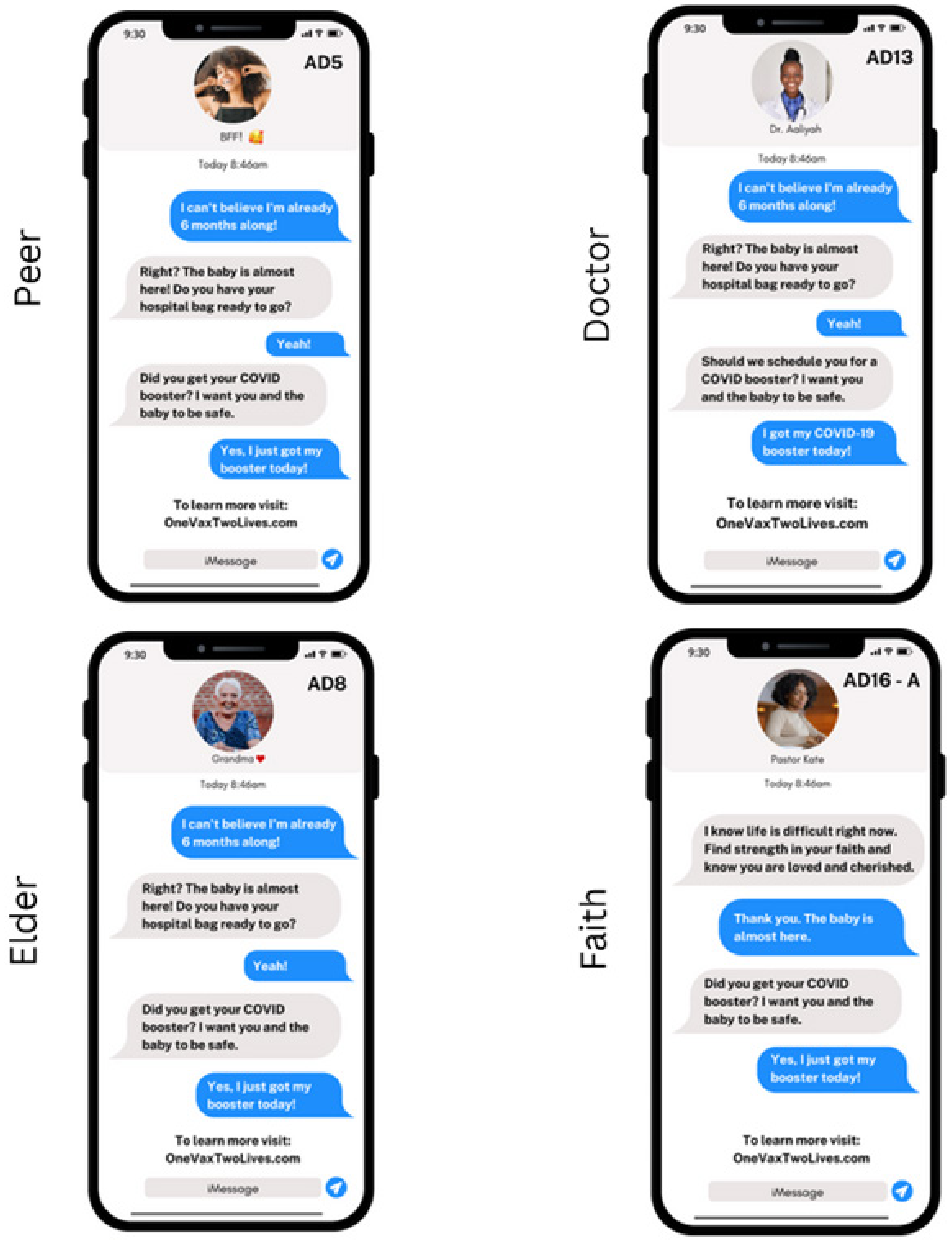

This figure depicts sample social media advertisements featuring an activation message, which are meant to provide an additional prompt to the viewer reminding them that they should get the COVID-19 vaccine.
